# The impact of hypoxia on B cells in COVID-19

**DOI:** 10.1101/2021.07.12.21260360

**Authors:** Prasanti Kotagiri, Federica Mescia, Aimee Hanson, Lorinda Turner, Laura Bergamaschi, Ana Peñalver, Nathan Richoz, Stephen D Moore, Brian M Ortmann, Benjamin J. Dunmore, Helene Ruffieux, Michael D Morgan, Zewen Kelvin Tuong, Rachael J M Bashford-Rogers, Myra Hosmillo, Stephen Baker, Anne Elmer, Ian G Goodfellow, Ravindra K. Gupta, Nathalie Kingston, Paul J. Lehner, Nicholas J. Matheson, Sylvia Richardson, Caroline Saunders, Michael P. Weekes, Cambridge Institute of Therapeutic Immunology and Infectious Disease-National Institute of Health Research (CITIID-NIHR) COVID BioResource Collaboration, Berthold Göttgens, Mark Toshner, Christoph Hess, Patrick. H. Maxwell, Menna. R. Clatworthy, James A. Nathan, John R. Bradley, Paul A. Lyons, Natalie Burrows, Kenneth G.C. Smith

## Abstract

Prominent early features of COVID-19 include severe, often clinically silent, hypoxia and a pronounced reduction in B cells, the latter important in defence against SARS-CoV-2. This brought to mind the phenotype of mice with VHL-deficient B cells, in which Hypoxia-Inducible Factors are constitutively active, suggesting hypoxia might drive B cell abnormalities in COVID-19. We demonstrated the breadth of early and persistent defects in B cell subsets in moderate/severe COVID-19, including reduced marginal zone-like, memory and transitional B cells, changes we also observed in B cell VHL-deficient mice. This was corroborated by hypoxia-related transcriptional changes in COVID-19 patients, and by similar B cell abnormalities in mice kept in hypoxic conditions, including reduced marginal zone and germinal center B cells. Thus hypoxia might contribute to B cell pathology in COVID-19, and in other hypoxic states. Through this mechanism it may impact on COVID-19 outcome, and be remediable through early oxygen therapy.

## Introduction

Hypoxemia is a prominent clinical feature of COVID-19. It often occurs silently, with many patients presenting to hospital in respiratory failure and with profoundly low blood oxygen saturations (Couzin-Frankel, 2020; Tobin, Laghi and Jubran, 2020). This asymptomatic, so-called “silent”, hypoxia is a particular trait of COVID-19 and associated with a worse outcome(Brouqui *et al*., 2021). Although poorly understood, it may arise from SARS-CoV-2 altering the respiratory drive, with blunted dyspnoea in the context of severe hypoxaemia(Tobin, Laghi and Jubran, 2020)(Gattinoni, Marini and Camporota, 2020)(Swenson, Ruoss and Swenson, 2021). Another early feature of moderate to severe COVID-19 is pronounced lymphopenia, with a marked reduction in many lymphocyte cell subsets usually maximal at presentation to hospital(Bergamaschi *et al*., 2021). The reduction in B cell subsets we observed in moderate/severe COVID-19 was reminiscent of the phenotype of mice with VHL-deficient B cells, which exhibit constitutive activation of Hypoxia-Inducible transcription Factors (HIFs)(Cho *et al*., 2016; Burrows *et al*., 2020), raising the possibility that hypoxia might contribute to B cell dysregulation in COVID-19. We therefore explored the possibility of a causal link between hypoxia and B cell abnormalities in COVID-19.

The B cell response is a vital component of immune defence against SARS-CoV-2. Neutralising antibodies contribute to protection from infection(Cao *et al*., 2020). In some contexts, monoclonal antibodies or convalescent serum can be of benefit(Katz, 2021). Patients with antibody deficiency are at increased risk of viral persistence(Buckland *et al*., 2020). Changes in B cell numbers are prominent in symptomatic COVID-19: increased plasmablasts and reduced memory B cells correlate with disease severity, and germinal centre (GC) responses, somatic hypermutation, and T follicular helper (TFH) cells may be reduced(Kaneko *et al*., 2020; Nielsen *et al*., 2020; Bergamaschi *et al*., 2021; Stephenson *et al*., 2021). Many of these changes persist beyond two months from symptom onset(Bergamaschi *et al*., 2021).

The marked abnormalities seen in the B cell response in moderate to severe COVID-19 may have a clinical impact. They might limit the speed and efficiency of the anti-SARS-CoV-2 response(Garcia-Beltran *et al*., 2021; Kemp *et al*., 2021) and predispose to reinfection or to secondary infection with different pathogens – the latter a major clinical problem in COVID-19(Zhou *et al*., 2020; Maes *et al*., 2021). Persistent B cell dysregulation also has the potential to contribute to other clinical sequelae of COVID-19, such as autoimmunity(Wang *et al*., 2020). Understanding the changes to B cell immunity caused by COVID-19, and the potential role of hypoxia as a mechanism underlying them, could therefore inform clinical management strategies.

Cells sense and respond to hypoxia through the two main HIF isoforms (HIF-1α and HIF-2α). When oxygen is present, these transcription factors are constitutively tagged for proteasomal degradation by prolyl-hydroxylase enzymes (PHDs)(Jaakkola *et al*., 2001), and the recruitment of the Von Hippel-Lindau (VHL) E3 ubiquitin ligase(Maxwell *et al*., 1999). In hypoxia, PHD activity is reduced, leading to HIF-α stabilisation, which modulates the expression of hundreds of genes(Schödel *et al*., 2011). During inflammation HIFs allow innate and adaptive immune cells to survive and function through regulation not only by hypoxia but also inflammatory stimuli such as Toll-like receptors (TLRs), and interleukin and antigen receptors(Clatworthy *et al*., 2014; Meng *et al*., 2018; Watts and Walmsley, 2019). B cell adaptation to hypoxia may be physiologically important, GCs are hypoxic(Cho *et al*., 2016; Burrows and Maxwell, 2017), and mice with B cell-specific VHL deletion and thus constitutively active HIF show abnormal B cell development and reduced GC B cells, antibody class-switching and affinity-maturation. Deleting HIF rescued the defects confirming a HIF-dependent effect (Cho *et al*., 2016; Burrows *et al*., 2020).

We explored the potential impact of reduced oxygen on B cells in detail. We first described the B cell changes associated with COVID-19 using data from a large prospective study, before performing a comparative analysis of these changes in mice in which B cell-specific VHL deficiency driven by either CD19-cre or MB1-cre. We then found evidence of a hypoxia transcriptional signature in early moderate to severe COVID-19, which single cell analysis demonstrated was particularly enriched in B cells. Finally, we explored the B cell immune response in mice housed in hypoxic conditions, allowing the impact of hypoxia on B cells to be examined in the absence of inflammation, something not possible in humans with COVID-19. This revealed that hypoxia could contribute to the marked B cell defects seen in COVID-19.

## Results

### Patient cohorts

SARS-CoV-2 PCR-positive subjects were recruited between 31^st^ March and 20^th^ July 2020 and divided into five categories according to peak clinical severity(Bergamaschi *et al*., 2021) (**Figure 1a**):

A. asymptomatic healthcare workers (HCWs) recruited from routine screening.
B. HCWs either still working with mild symptoms, or symptomatic and self-isolating.
C. patients who presented to hospital but never required oxygen supplementation.
D. admitted patients whose maximal respiratory support was supplemental oxygen.
E. patients who required assisted ventilation (57) or died without ventilation (3).

**Figure 1.**
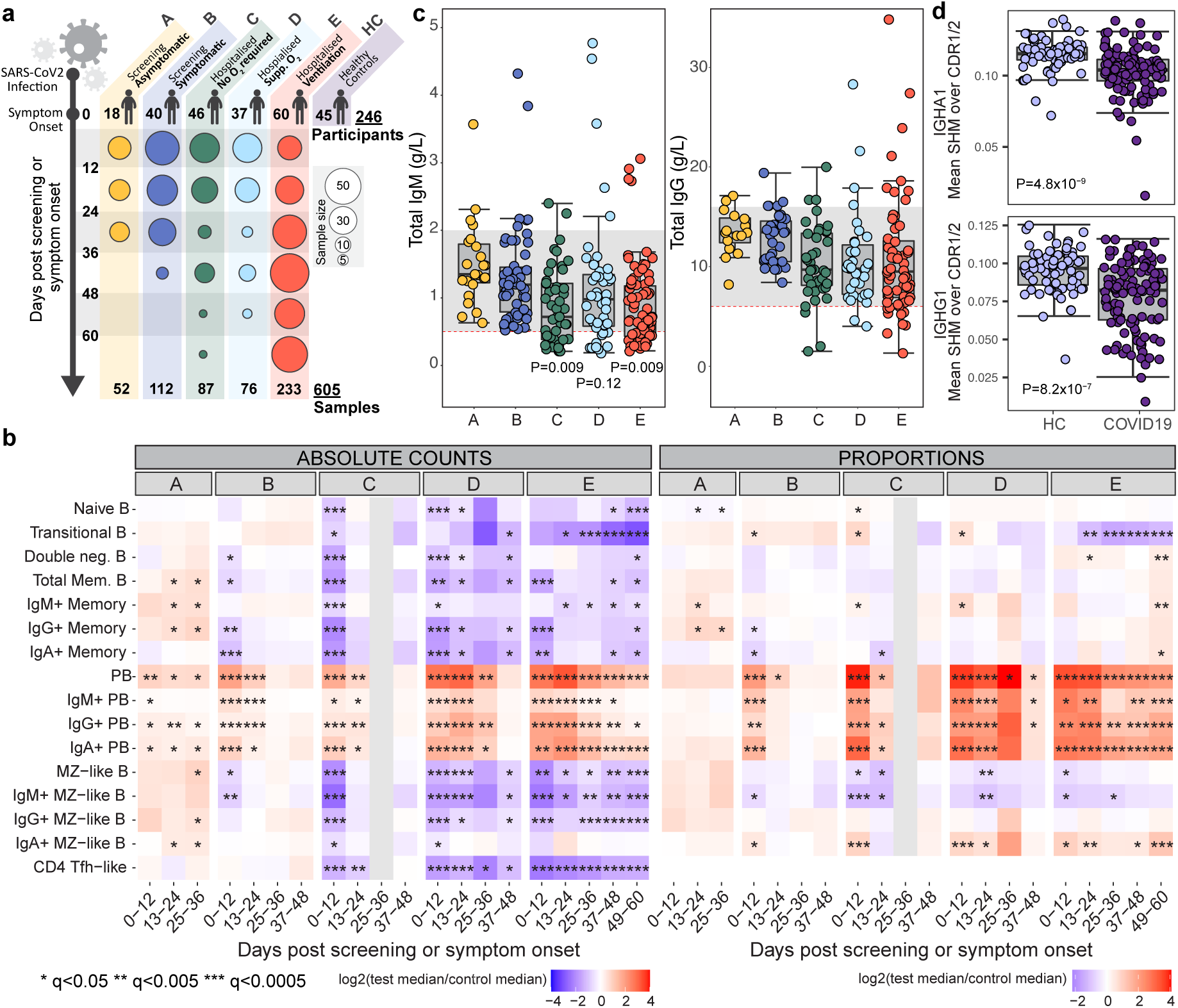
B cells in COVID-19. **a**, Cohort details. Time post positive swab (group A) or symptom onset (groups B-E). **b**, Median absolute cell counts (left) or proportions relative to total B cells (right) (log2 fold change relative to healthy controls). Wilcoxon test FDR adjusted p-value (q): *<0.05, **<0.005, ***<0.0005. **c**, Serum IgG and IgM (g/L) at enrolment. Grey band: 5-95^th^ centiles of healthy reference range (see methods). Significant P values listed. **d**, Somatic hypermutation frequency in IgA and IgG within 0-12 days post symptom onset, calculated over the CDR1/CDR2 regions using BCR sequencing of whole blood. Wilcoxon test p-value. **c**,**d**, Circles represent individual donors.

Patients were bled approximately weekly while inpatients, and less frequently after discharge. Patient courses are measured in time since symptom onset for groups B to E, and from their first positive swab for group A (as they are asymptomatic, leading to them being sampled, on average, later post-infection than groups B-E).

### Persistent B cell dysregulation in COVID-19

We compared absolute B cell subset numbers in COVID-19 patients to 45 healthy controls, including some previously reported data for comparison (**Figure 1b**)(Bergamaschi *et al*., 2021). In asymptomatic HCWs (group A) an often non-significant increase in most B cell subsets occurred, while in Group B mild and short-lived reductions in IgA and IgG memory and in marginal zone-like (MZL) B cells were seen. Plasmablasts in both groups were increased early before declining slowly. In contrast, in all more severe groups (C-E), profound reductions in many B cell subsets were seen at the first bleed, including naïve and transitional B cells, and memory and MZL B cells of all isotypes. Most subsets then showed some recovery, though transitional B cells continued to fall progressively in severe disease. T_FH_-like CD4 T cells were reduced out to 60 days in groups D and E (**Figure 1b**). Cell subsets are also shown as a proportion of total B cells, which detected the increase in plasmablasts, but underestimated or missed most other changes (**Figure 1b**), explaining why most studies fail to fully appreciate the profound B cell dysregulation seen in COVID-19. Single-cell RNA-sequencing coupled with analysis of surface proteins on a subset of patients confirmed proportional differences (**Figure S1**).

Total serum IgM fell as disease severity increased, with many patients in groups C-E having IgM levels below the normal range, while IgG and IgA were less impacted (**Figures 1c and S1a**). Anti-SARS-CoV-2 spike antibodies rose over time in all severity groups, reaching highest titres in the more severe groups(Bergamaschi *et al*., 2021). BCR sequencing showed reduced somatic mutation in COVID-19 patients, most prominent in IgA and IgG1/2 (**Figures 1d and S1b**), consistent with previous reports(Nielsen *et al*., 2020).

Having observed the similarity in B cell phenotype in COVID-19 and mice with constitutively active HIF (Cho *et al*., 2016; Burrows *et al*., 2020), we hypothesised that the loss of B cells might be related to hypoxia *in vivo*. While some *in vitro* hypoxic studies on B cells have been performed (Cho et al., 2016; Burrows et al., 2020), the effects of acute hypoxia on immune responses in mice and humans have not been assessed, which is what we addressed in this study. Immunised *Vhl*^*-/-*^*Cd19-cre* mice (in which VHL is deleted at the pre-B cell stage) were then studied to allow more accurate comparison with the B cell pathology in COVID-19: they showed reductions in splenic follicular and MZ B cells (consistent with observations in unimmunised mice(Xu *et al*., 2019). We further observed reduced GC B cells, and an increased plasma cell (PC) to B cell ratio **(Figures 2a and S2a)**. Findings were confirmed in *Vhl*^*-/-*^*Mb1-cre* mice (VHL deleted in pro-B cells) immunised with NP-KLH, in which reduced NP-specific GC and memory B cells, and TFH cells were also observed. Marked reductions were observed in transitional B cells, consistent with observations in unimmunised mice(Burrows *et al*., 2020). Changes were more pronounced in spleen than lymph node (**Figures 2b and S2b-d**).

**Figure 2.**
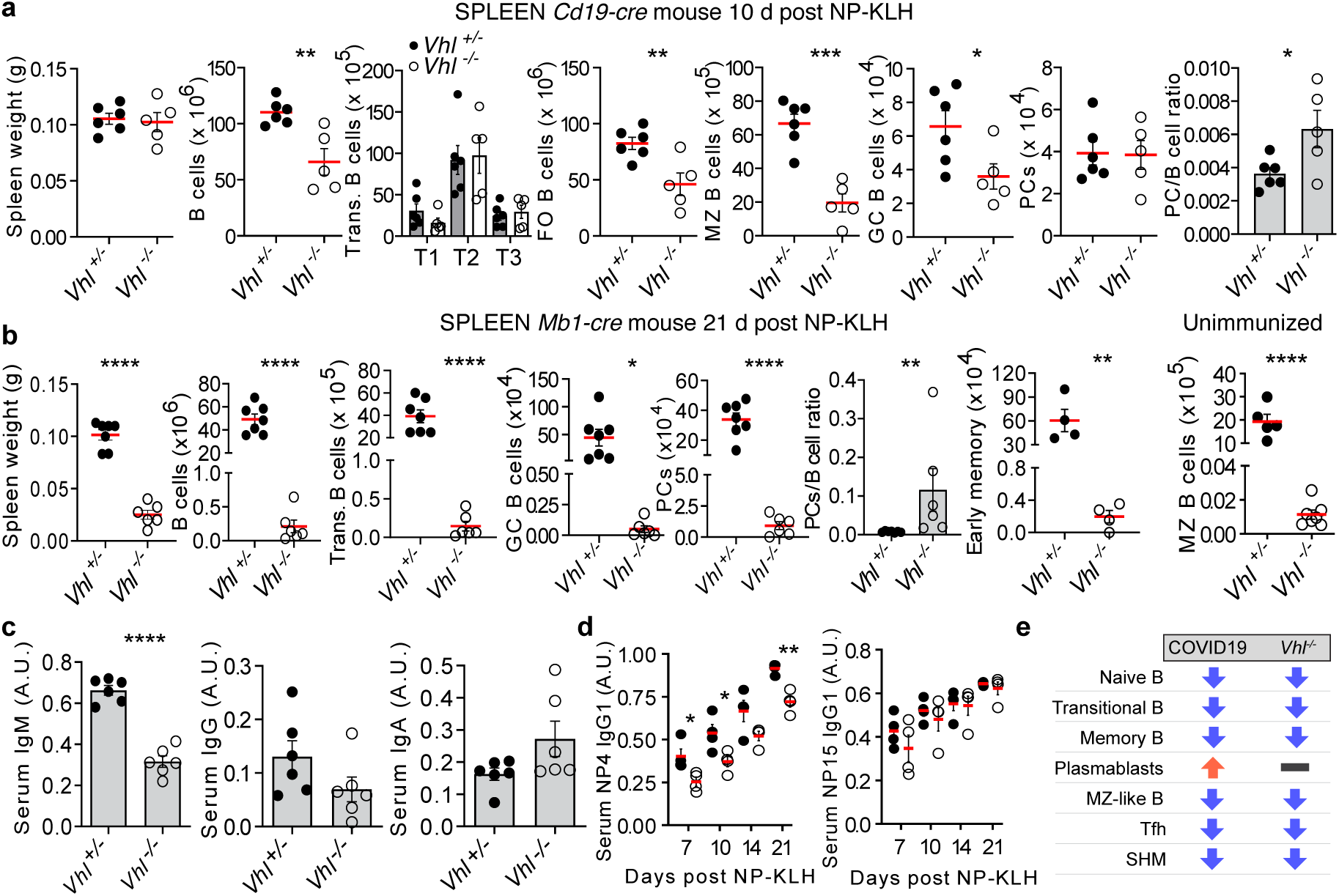
B cells in VHL-deficient mice. **a**, Weight and B cell flow cytometric data from *Vhl*^+/-^*Cd19-cre* and *Vhl*^-/-^*Cd19-cre* mice 10 days post NP-KLH immunization; Spleen B cells (B220^+^), Transitional (T1–T3: B220^+^CD93^+^CD23 IgM), FO (B220^+^CD93^−^CD23^+^CD21^+^), MZ (B220^+^CD93^−^CD23^−^CD21^+^), GC B cells (B220^+^CD95^high^GL-7^high^) and PCs (B220^-^CD138^+^). \**P* < 0.05,\*\**P* < 0.01, \*\*\**P* < 0.001, unpaired, two-sided Student’s t-test. **b**, Weight and B cell flow cytometric data from *Vhl*^+/-^*Mb1-cre* and *Vhl*^-/-^*Mb1-cre* mice 21 days post NP-KLH immunization in spleen; B cells, transitional, FO, GC, PC and early memory B cells (B220^+^IgD^neg/lo^CD95^+^GL7^-^CD38^+^CD73^+^) and PC:B cell ratio displayed. PCs were not increased in absolute number, but the PC:B cell ratio was consistently increased. MZ B cells are from unimmunized mice. \**P* < 0.05,\*\**P* < 0.01, \*\*\*\**P* < 0.0001, unpaired, two-sided Student’s t-test. **c**, Serum IgM, IgG and IgA in naïve *Vhl*^+/-^*Mb1-cre* and *Vhl*^-/-^*Mb1-cre* mice, by ELISA (A.U., arbitrary units). Unpaired, two-sided Student’s t-test. **d**, Serum NP-specific antibody titres after NP-KLH immunization in *Vhl*^+/-^*Mb1-cre* and *Vhl*^-/-^*Mb1-cre* mice, by ELISA. two-way ANOVA with Sidak’s multiple comparisons test. **a-d**, mean ± s.e.m, individual mice shown, each of 1 experiment representative of between 2 and 9. \**P* < 0.05,\*\**P* < 0.01,\*\*\**P* < 0.001, \*\*\*\**P* < 0.0001 **e**, B cell phenotype comparison in patients with COVID-19 and *Vhl*^-/-^*Mb1-cre* mice.

Serum IgM, but not IgG and IgA, was reduced, as was affinity maturation (**Figure 2c and d**) and somatic hypermutation (SHM) in some isotypes (**Figure S2e**). Reduced GC and memory B cells, along with defects in affinity maturation, were similar to those observed in an inducible model of B cell specific *Vhl* deletion(Cho *et al*., 2016). Together, these observations highlight that HIF has robust and consistent impacts on B cells across multiple models of genetic HIF activation, and that the reductions in B cells seen are unlikely due solely to defects in B cell development, as they occur rapidly in inducible as well as constitutive models. This evidence demonstrates that the B cell dysregulation in mice with B cell-specific VHL-deletion was remarkably similar to that seen in patients with moderate to severe COVID-19 (**Figure 2e**), supporting the possibility that signalling through HIF could be implicated in the B cell abnormalities seen in COVID-19.

### Early transcriptional changes characteristic of hypoxia correlate with B cell changes in COVID-19

Hypoxia in COVID-19 patients, as determined by monitoring peripheral oxygen saturation (SpO_2_), was common early in disease and tended to improve with recovery, or with ventilation or ECMO in intensive care (group E) (**Figure S1d**), but was hard to correlate directly with immune changes, as recorded SpO_2_ is usually taken on oxygen replacement, which is commonly administered in the ambulance or immediately on arrival in hospital. Thus these data will underestimate the real hypoxia on admission, which is likely to have been sustained for hours or days before the patient presents. Furthermore, SpO_2_ is not reliably reflective of tissue hypoxia, and the provision of oxygen supplementation was variable and its documentation hard to quality control. We therefore instead measured the impact of hypoxia on the transcriptome in whole blood and single cells. We used singular value decomposition to calculate an eigengene representative of the curated Hallmark hypoxia signature within the whole blood dataset and showed that this was associated with disease severity. Eigengene values were similar to health in groups A and B, and rose with increasing severity in groups C, D and E (**Figure 3a**). Gene set enrichment analysis (GSEA) of the Hallmark hypoxia geneset showed association with disease severity, in particular group E early, and group C later, perhaps reflecting severity and lack of O_2_ supplementation respectively (**Figure 3b** and **S1e**).

**Figure 3.**
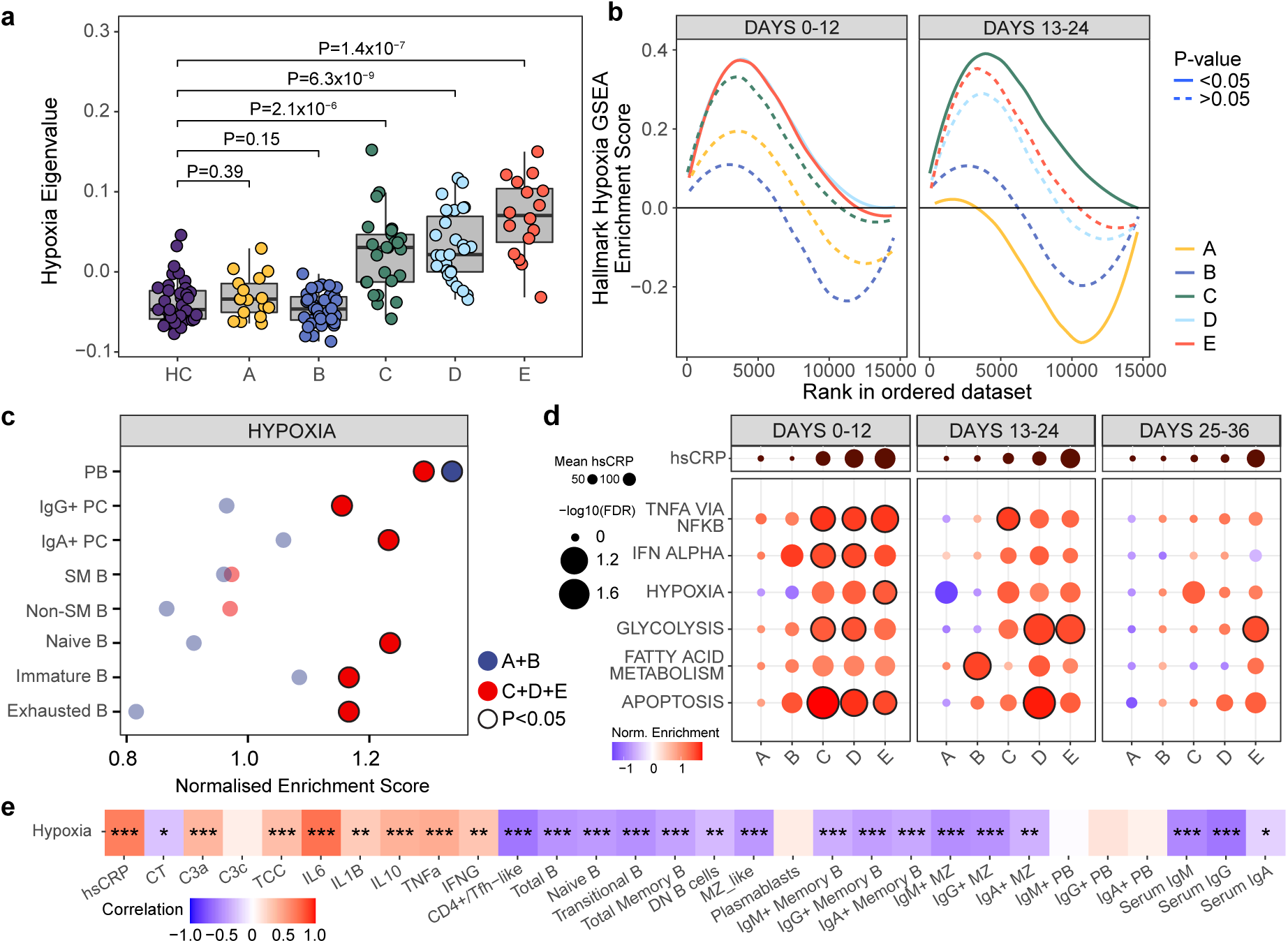
Hypoxia-related transcription signatures and COVID-19. **a**, Eigenvalues of Hallmark Hypoxia geneset grouped by severity at 0-12 days post symptom onset, unpaired, two-sided Student’s t-test. Circles represent individual donors. **b**, GSEA of Hallmark Hypoxia geneset in COVID-19 versus HC grouped by severity and time, Kolmogorov-Smirnov statistic applied. **c**, B cell subpopulations identified using CITEseq with GSEA of Hallmark Hypoxia geneset assessed on a single cell level comparing HC to COVID-19, grouped by severity. Samples within 24 days of symptom onset (B-E)/positive swab (A). Severity groups A/B n=8, C/D/E n =20. Outlined circles represent P <0.05. **d**, GSEA assessing enrichment of Hallmark genesets in COVID-19 versus HC grouped by severity and 12-day time bins, FDR adjusted p-value. Outlined circles represent a nominal P value <0.05 and FDR adjusted P <0.2. Mean hsCRP represented. **e**, Correlation between Hallmark hypoxia geneset eigengenes and absolute cell counts, complement levels, select cytokines and total serum immunoglobulins, at 0-12 days post symptom onset in COVID-19 patients. Boxes coloured by strength of correlation, Pearson correlation.

To explore the impact of hypoxia on specific B cell subsets, we looked for enrichment of this Hallmark gene set across B cell subtypes identified by single-cell RNA-sequencing(Stephenson *et al*., 2021). This showed a tendency for enrichment in groups C, D and E in all cell types apart from memory B cells. Patients in groups A and B showed such enrichment only in plasma cells (**Figure 3c**).

The “Hallmark hypoxia” signature was enriched for genes regulated by HIF, and could therefore be activated by both reduced oxygen-tension or inflammatory stimuli. We therefore compared Hallmark signatures of hypoxia with inflammation in whole blood - the hypoxia signature was prominent in early severe disease (groups C-E) before declining, perhaps due to recovering disease and effective oxygen supplementation, but was not enriched in mild disease (A and B). In contrast inflammation-related signatures were often seen in these mild groups, but were also prominent early in severe disease (**Figure 3d**). The differential enrichment of hypoxic and inflammatory signatures in asymptomatic and mild disease suggests a specific role for hypoxia, but an additional role for inflammation cannot be excluded (**Figure 3d**). Finally, we found that the hypoxia eigengene correlated inversely with B cell number across most subsets, with the exception of plasma cells (see discussion) (**Figure 3e**).

### Reduced MZ, GC and transitional B cells in hypoxic wild-type mice

The findings in mice with VHL-deficient B cells provided evidence that HIF-regulated gene activation can contribute to B cell defects of the sort seen in COVID-19, and so, in order to differentiate inflammatory-driven from hypoxia-driven HIF-mediated effects, we studied mice in hypoxic conditions (10% O_2_) after immunisation with NP-KLH. After 11 days of hypoxia we observed reduced transitional, FO, MZ and GC B cells, whilst PCs were normal (**Figure S3a-b)**. These defects persisted when hypoxia was prolonged (20 days; **Figure 4a-b and S3c)**. In hypoxic mice there was a tendency to reduced early memory B cells **(Figure 4a and S3d)** and serum Ig was normal but antigen-specific IgG1 was reduced **(Figure 4c and S3e)**. Histological analysis revealed that, in hypoxic conditions, B cells were almost absent from the MZ, which appeared otherwise structurally intact **(Figure 4d and S3f-g)**. Hypoxia induced only minor reductions in T cells and NK cells, and no changes in macrophage numbers, indicating that B cells seem particularly sensitivity to perturbations in oxygenation and HIF **(Figure S4 and data not shown)**. Thus while it is likely that hypoxia will have other effects, that will warrant more detailed examination, B cells seem particularly sensitivity to perturbations in oxygenation and HIF activity. Some mice were removed from the hypoxic chamber after 11 days: B cell subsets generally recovered following this reoxygenation. MZ and GC B cells were most prominently affected by hypoxia, continuing to decline under hypoxic conditions, and recovering more slowly following reoxygenation, than other subsets **(Figure 4e)**.

**Figure 4.**
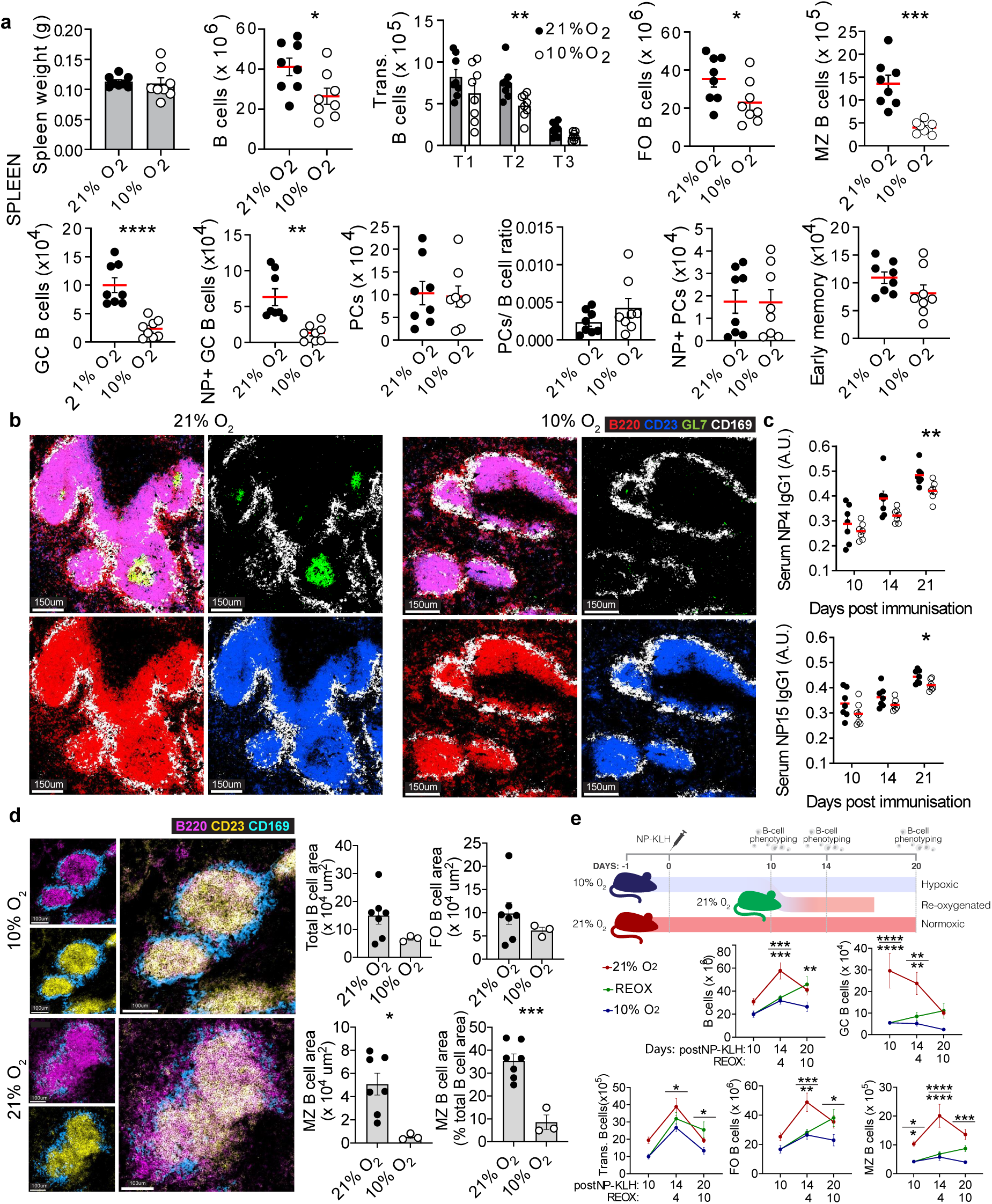
The response of mouse B cells to hypoxia *in vivo*. **a**, WT mice were exposed to 21% O_2_ (normoxia) or 10% O_2_ (hypoxia), were immunized with NP-KLH at d 1, then at d 21 spleen B cell subsets were enumerated by flow cytometry. Transitional, FO, MZ, GC B cells (gated as in Fig. 2a), early memory B cells (B220^+^IgD^neg/lo^CD95^+^GL7^-^CD38^+^CD73^+^) and PCs (B220^-^CD138^+^). \**P* < 0.05,\*\**P* < 0.01,\*\*\**P* < 0.001 unpaired, two-sided Student’s t-test. FO, \**P* < 0.05 two-sided Mann-Whitney U-test. **b**, Representative spleen confocal images from immunised mice described in a. GC B cells (green, B220^+^GL7^+^), B cells (red, B220^+^), FO B cells (blue, B220^+^CD23^+^) and MZ metallophilic macrophages (white, CD169^+^). **c**, Serum NP-specific antibody titres after NP-KLH immunization in normoxic and hypoxic mice. \**P* < 0.05,\*\**P* < 0.01 unpaired, two-sided Student’s t-test. **d**, Representative spleen confocal images and B cell and MZ area quantification from immunised mice described in a. MZ B cells (pink, B220^+^CD23^-^), FO B cells (yellow, B220^+^CD23^+^) and MZ metallophillic macrophages (blue, CD169^+^). Symbols represent individual follicles from one spleen per condition. \**P* < 0.05,\*\*\**P* < 0.001 unpaired, two-sided Student’s t-test. **e**, Experiment outline and absolute numbers of total, transitional, FO, MZ and GC B cells by flow cytometry, from spleens. \**P* < 0.05,\*\**P* < 0.01,\*\*\**P* < 0.001,**** *P* < 0.0001 two-way ANOVA with Tukey’s multiple comparisons test. **(a**,**c)** *n* = 8 21% O_2_ and 8 10% O_2_, **(e)** 8 mice per group. **(a**,**c and e)** Mean ± s.e.m., data pooled from two independent experiments, results confirmed in a third.

## Discussion

Our study demonstrates profound B cell abnormalities in patients with severe COVID-19, and provides evidence that they may be largely driven by hypoxia. B cell lymphopenia extends across all subsets, is present soon after symptom onset, and is often persistent. There is an associated reduction in total serum IgM and in somatic hypermutation in switched B cells. The exceptions are plasmablasts, which rise early irrespective of severity, and remain elevated in severe COVID-19. Despite these early changes, patients in all groups develop neutralising anti-SARS-CoV-2 antibodies(Bergamaschi *et al*., 2021), which doesn’t require affinity maturation(Clark *et al*., 2021). It nonetheless seems likely that these profound B cell deficits could have an impact on disease, perhaps in part because affinity maturation might be required to generate a broader spectrum of neutralizing antibodies as disease progresses(Clark *et al*., 2021), which might, for example, help prevent re-infection by new variant strains of virus. B cell dysregulation might also play a part in driving the autoimmunity which is increasingly thought to be important in COVID-19(Wang *et al*., 2020).

Bystander CD8 T cell responses are delayed in severe COVID-19, perhaps reducing early viral control(Bergamaschi *et al*., 2021). Defects in non-antigen specific B cell function may play a similar role, for example in reducing early antigen localisation, transport or presentation, or cytokine or “natural” antibody production(Hernandez and Holodick, 2017). Later in the disease course B cell defects may predispose to secondary bacterial infection, which is a major clinical problem in severe COVID(Maes *et al*., 2021; Ripa *et al*., 2021). Marginal zone B cells are key players in early defence against blood-borne bacterial infection(Nemazee, 2021) and their dysregulation may play a role in autoimmunity (Tull et al., 2021). MZL B cells are profoundly reduced in COVID-19 in humans, and hypoxia results in MZs almost devoid of B cells in mice. MZL B cells in humans, and MZ cells in hypoxic mice, also recover more slowly than other B cell subsets. The reasons for this are unknown, but human MZ B cells appear to be derived from T2 transitional B cells and/or memory B cells (Kibler *et al*., 2021; Tull *et al*., 2021), both of which are profoundly depleted in COVID-19. This alone may hamper recovery, but additional unknown factors may also determine reconstitution. The loss of GCs in the spleen and lymph nodes has been reported in post-mortem studies of COVID-19 patients (Kaneko *et al*., 2020), consistent with the reduced somatic hypermutation seen in human COVID-19(Figure 1d and Nielsen et al., 2020), and with our observations in hypoxic mice. Hypoxia plays a role in controlling GC responses and can increase apoptosis (Cho et al., 2016), and this may be directly responsible for their loss. GC recovery is likely to be governed by the regeneration of their precursors, and presumably antigen-driven expansion. These defects in both MZ and GC B cells suggests hypoxia may directly contribute to the risk of secondary infection in COVID-19, and perhaps in other hypoxia-associated clinical situations. Determination of the extent of depletion of these cells in human tissue, and their rate of recovery, might guide strategies to prevent such sequelae.

In COVID-19, HIF is likely to be activated by hypoxia, inflammation and antigen receptor/BCR cross-linking(Meng *et al*., 2018). There is a tighter correlation between disease severity and hypoxia than that observed with inflammatory signatures, but nevertheless it is impossible to conclusively separate the two impacts. Hypoxia exerts its major effect on HIF-1α by preventing its degradation, while the main inflammatory impact is through enhancing HIF transcription, making a synergistic impact of hypoxia and inflammation on HIF function plausible(Burrows and Maxwell, 2017; Watts and Walmsley, 2019). Our demonstration that hypoxia alone can induce profound, reversible B cell abnormalities in mice is novel, and supports a role for hypoxia in driving B cell abnormalities in COVID-19. T cell numbers were minimally impacted by hypoxia, suggesting that, while the impact of hypoxia on other components of the immune response warrants more detailed examination, its effect seems relatively B cell specific.

HIF regulation might be different in plasmablasts. In groups A and B, where hypoxia does not occur, plasmablast numbers are increased and they are the only subset in which HIF-related transcriptional signatures are enriched. Plasmablasts are the only B cell subset in which cell numbers do not correlate with the hypoxia signature, and their numbers are stable in mice subject to hypoxia. Thus, plasmablasts appear to show signs of HIF activation in the absence of hypoxia, raising the possibility that HIF is constitutively active in these cells. This is consistent with a growing literature demonstrating that HIF-1α is active in multiple myeloma(Martin *et al*., 2011). Given this aspect of COVID-19 B cell pathology does not appear impacted by hypoxia, therapeutic approaches to it may need to involve pharmacological antagonism of HIF.

The observation that hypoxia perturbs B cell immunity has implications in a wide range of clinical settings. In COVID-19 these observations lead to the prediction that early and aggressive oxygen therapy may lead to improved immune responsiveness and thus clinical outcome.

## Methods

### Participant recruitment and clinical data collection

This cohort has been previously described by Bergamaschi et al.(Bergamaschi *et al*., 2021) Briefly, study participants were recruited between 31/3/2020 and 20/7/2020 from patients attending Addenbrooke’s Hospital, Royal Papworth Hospital NHS Foundation Trust or Cambridge and Peterborough Foundation Trust with a confirmed diagnosis of COVID-19, together with Health Care Workers identified through staff screening as PCR positive for SARS-CoV-2(Rivett *et al*., 2020). Controls were recruited among hospital staff attending Addenbrooke’s for SARS-CoV-2 serology screening programme and having a negative serology result. Ethical approval was obtained from the East of England – Cambridge Central Research Ethics Committee (“NIHR BioResource” REC ref 17/EE/0025, and “Genetic variation AND Altered Leucocyte Function in health and disease - GANDALF” REC ref 08/H0308/176). All participants provided informed consent.

Inpatients were sampled at study entry, and then at regular intervals as long as they remained admitted to hospital (approximately weekly up to 4 weeks, and then every 2 weeks up to 12 weeks). Discharged patients were invited to provide a follow-up sample 4-8 weeks after study enrolment. Health care workers were sampled at study entry, and subsequently after approximately 2 and 4 weeks.

Clinical data were retrospectively collected by review of medical charts and extraction of data (laboratory test results, vital signs, medications) from Epic electronic health records (Addenbrooke’s Hospital) and from MetaVision ICU (Royal Papworth Hospital).

Study volunteers were classified in 5 groups:

- Group A: health care workers who were asymptomatic at the time of positive SARS-CoV-2 testing. This group included 10 volunteers who had possible COVID-19 symptoms before PCR testing (median time from symptoms to COVID-19 PCR test 26 days, range 9-42 days).
- Group B: health care workers who had possible COVID-19 symptoms at the time of PCR testing.
- Group C: patients in hospital who did not receive any supplemental oxygen for COVID-19. Five patients were discharged soon after initial diagnosis and assessment but followed up as part of the study.
- Group D: patients in hospital who received supplemental oxygen using low flow nasal prongs, simple face mask, Venturi mask or non re-breather face mask
- Group E: patients in hospital who received any of non-invasive ventilation (NIV), mechanical ventilation or ECMO. Patients who received supplemental oxygen (but no ventilation) and deceased in hospital were also assigned to group E.

Study results were analysed according to time since onset of COVID-19 symptoms, or otherwise time since positive SARS-CoV-2 testing (in group A and in 4 asymptomatic patients in group C).

### Peripheral blood mononuclear cell preparation and flow cytometry immunophenotyping

For direct enumeration of T, B and NK cells, an aliquot of whole blood EDTA (50μl) was added to BD TruCount™ tubes with 20μl BD Multitest™ 6-colour TBNK reagent (BD Biosciences) and processed as per the manufacturer’s instructions.

Peripheral venous blood (up to 27 ml per sample) for isolation of Peripheral Blood Mononuclear Cells (PBMCs) was collected into 10% sodium citrate tubes. PBMCs were isolated using Leucosep tubes (Greiner Bio-One) with Histopaque 1077 (Sigma) by centrifugation at 800x g for 15 minutes at room temperature. PBMCs at the interface were collected, rinsed twice with autoMACS running buffer (Miltenyi Biotech) and cryopreserved in FBS with 10% DMSO. All samples were processed within 4 hours of collection.

Approximately 10^6^ cells have been stained with: anti-human IgM (clone: G20-127, BD), CD19 (clone: SJ25C1, BD), CD38 (clone: HIT2, BD), IgD (clone: IA6-2, BD), CD20 (clone: 2H7, BD), CD3 (clone: UCHT1, BioLegend), CD14 (clone: 63D3, BioLegend), CD15 (clone: W6D3, BioLegend), CD193 (clone: 5E8, BioLegend), CD27 (clone: O323, BioLegend), CD56 (clone: MEM188, Thermo), CD24 (clone: ML5, BD), IgA (polyclonal goat IgG, Jackson), IgG (clone: G18-145, BD), and Zombie Yellow (BioLegend) as described in detail by Bergamaschi et al.(Bergamaschi *et al*., 2021) Samples were stored at 4°C and acquired within 4 hours using a 5-laser BD Symphony X-50 flow cytometer. Single colour compensation tubes (BD CompBeads) or cells were prepared for each of the fluorophores used and acquired at the start of each flow cytometer run.

Samples were gated in FlowJo v10.2 and number of cells falling within each gate was recorded. For analysis, these were expressed either as proportion of total B cells or an absolute concentration of cells per μl, calculated using the proportions of daughter populations present within the parent population determined using the BD TruCount™ system.

### CRP, complement components and cytokines

As detailed in Bergamaschi et al.(Bergamaschi *et al*., 2021), concentrations of complement components were measured in EDTA plasma using commercially available enzyme-linked immunosorbent assays (ELISA) kits. High sensitivity CRP and cytokines (IL-6, IL-10, IL-1β, TNFα and IFNγ) were assayed in serum using standard laboratory assays.

### Total Immunoglobulin levels

Serum immunoglobulin levels were measured for 186 COVID-19 patients and 45 healthy controls at the time of enrolment using the standard assay by the Immunology Department at Peterborough City hospital.

### Whole blood bulk RNA-Seq

Whole blood RNA was extracted from PAXgene Blood RNA tubes (BD Biosciences) of 188 COVID-19 patients at up to 2 time points and 42 healthy volunteers. RNA-Sequencing libraries were generated using the SMARTer® Stranded Total RNA-Seq v2 - Pico Input Mammalian kit (Takara) using 10ng RNA as input following the manufacturer’s protocol. Libraries were pooled together (n = 96) and sequenced using 75bp paired-end chemistry across 4 lanes of a Hiseq4000 instrument (Illumina) to achieve 10 million reads per sample. Read quality was assessed using FastQC v.0.11.8 (Babraham Bioinformatics, UK), and SMARTer adaptors trimmed and residual rRNA reads depleted in silico using Trim galore v.0.6.4 (Babraham Bioinformatics, UK) and BBSplit (BBMap v.38.67(BBMap - Bushnell B. - sourceforge.net/projects/bbmap/)), respectively. Alignment was performed using HISAT2 v.2.1.0 (Kim et al., 2019) against the GRCh38 genome achieving a greater than 95% alignment rate. Count matrices were generated using featureCounts (Rsubreads package)(Liao, Smyth and Shi, 2019) and stored as a DGEList object (EdgeR package)(Robinson, McCarthy and Smyth, 2009) for further analysis.

All downstream data handling was performed in R (R Core Team, 2015). Counts were filtered using filterByExpr (EdgeR) with a gene count threshold of 10 CPM and the minimum number of samples set at the size of the smallest disease group. Library counts were normalised using calcNormFactors (EdgeR) using the method ‘weighted trimmed mean of M-values’. The function ‘voom’(Law *et al*., 2014) was applied to the data to estimate the mean-variance relationship, allowing adjustment for heteroscedasticity.

The analyses were carried out splitting the samples in 12 days bins post screening (group A) or symptom onset (groups B-E) as described in supplementary table 1.

### Single cell RNA-seq

CITE-seq data were generated from frozen PBMCs of 36 COVID-19 patients and 11 healthy controls as described by Stephenson et al.(Stephenson *et al*., 2021) Briefly, after thawing, pools of 4 samples were generated by combined 500,000 viable cells per individual (total of 2 million cells per pool). TotalSeq-C™ antibody cocktail (BioLegend 99813) was used to perform cell surface marker staining on 500,000 cells per pool. 50,000 live cells (up to a maximum of 60,000 total cells) for each pool were processed using Single Cell V(D)J 5’ version 1.1 (1000020) together with Single Cell 5’ Feature Barcode library kit (1000080), Single Cell V(D)J Enrichment Kit, Human B Cells (1000016) and Single Cell V(D)J Enrichment Kit, Human T Cells (1000005) (10xGenomics) according to the manufacturer’s protocols. Samples were sequenced on NovaSeq 6000 (Illumina) using S1 flowcells. Droplet libraries were processed using Cellranger v4.0. Reads were aligned to the GRCh38 human genome concatenated to the SARS-Cov-2 genome (NCBI SARS-CoV-2 isolate Wuhan-Hu-1) using STAR(Dobin *et al*., 2013) and unique molecular identifiers (UMIs) deduplicated. CITE-seq UMIs were counted for GEX and ADT libraries simultaneously to generate feature X droplet UMI count matrices.

### Statistics

All statistical analyses were conducted using custom scripts in R (R Core Team, 2015). Absolute cell counts (cells/uL) were offset by +1 to allow subsequent log2 transformation of zero counts. Unless otherwise specified, longitudinally collected data was grouped by bins of 12 days from symptom onset or first positive SARS-CoV2 swab. Pairwise statistical comparisons of absolute cell counts and proportions and immunoglobulin levels between individuals in a given severity group at a given time bin and HCs, or between severity groups, was conducted by Wilcoxon test unless otherwise specified. For analyses involving repeated measures, false discovery rate corrected (Benjamini & Hochberg) p values were reported. For individuals sampled more than once within a given time bin, data from the earliest blood collection was used.

Gene set enrichment analysis (GSEA)(Subramanian *et al*., 2005) was used to identify biological pathways enriched in COVID-19 severity groups relative to healthy controls. Briefly, a list of ranked genes, determined by Signal-To-Noise ratio was generated. An enrichment score was calculated, determined by how often genes from the geneset of interest appeared at the top or the bottom of the pre-ranked set of genes with the enrichment score representing the maximum deviation from zero. To assess statistical significance, an empirical phenotype-based permutation test was run, where a collection of enrichment scores was generated from the random assignment of phenotype to samples and used to generate a null distribution. To account for multiple testing, an FDR rate q < 0.20 was deemed significant. A leading edge analysis was performed to determine the genes contributing the most to the enrichment of a given pathway and was subsequently illustrated in a heatmap. HALLMARK gene sets from the Molecular Signatures Database (http://www.broadinstitute.org/gsea/msigdb) were used in analysis.

The relationships between immunological parameters and transcriptional data in the form of gene expression modules were assessed using Pearson’s correlation (Hmisc package) and visualized with corrplot.

### B Cell Receptor Repertoire

#### Library Preparation

B cell receptor repertoire libraries have been generated for 119 COVID-19 patients and 37 healthy controls using the protocol describe by Bashford-Rogers et al.(Bashford-Rogers *et al*., 2019). Briefly, 200ng of total RNA from PAXgenes (14ul volume) was combined with 1uL 10mM dNTP and 10uM reverse primer mix (2uL) and incubated for 5 min at 70°C. The mixture was immediately placed on ice for 1 minute and then subsequently combined with 1uL DTT (0.1 M), 1uL SuperScriptIV (Thermo Fisher Scientific), 4ul SSIV Buffer (Thermo Fisher Scientific) and 1uL RNAse inhibitor. The solution was incubated at 50 °C for 60 min followed by 15 min inactivation at 70 °C. cDNA was cleaned with AMPure XP beads and PCR-amplified with a 5′ V-gene multiplex primer mix and 3′ universal reverse primer using the KAPA protocol and the following thermal cycling conditions: 1cycle (95°C, 5min); 5cycles (98°C, 20s; 72°C, 30s); 5cycles (98°C, 15s; 65°C, 30s; 72°C, 30s); 19cycles (98 °C, 15s; 60°C, 30s; 72°C, 30s); 1 step (72°C, 5 min). Sequencing libraries were prepared using Illumina protocols and sequenced using 300-bp paired-end sequencing on a MiSeq.

#### Sequence analysis

Raw reads were filtered for base quality using a median Phred score of ≥32 (http://sourceforge.net/projects/quasr/). Forward and reverse reads were merged where a minimum 20bp identical overlapping region was present. Sequences were retained where over 80% base sequence similarity was present between all sequences with the same barcode. The constant-region allele with highest sequence similarity was identified by 10-mer matching to the reference constant-region genes from the IMGT database. Sequences without complete reading frames and non-immunoglobulin sequences were removed and only reads with significant similarity to reference IGHV and J genes from the IMGT database using BLAST were retained. Immunoglobulin gene use and sequence annotation were performed in IMGT V-QUEST, and repertoire differences were performed by custom scripts in Python.

### Animal Models

#### Mice

*Vhl*^−/−^ mice(Haase *et al*., 2001) were crossed with *Cd79a-cre* (*Mb1-cre*)(Hobeika *et al*., 2006) or *Cd19-cre* (JAX, stock no. 004126) to delete *Vhl* in the B cell lineage. All mice with *lox*P-flanked alleles were hemizygous for *Cre*. Deletion efficiency was determined via real-time PCR of genomic DNA. The degree of excision was calculated by comparison of *Vhl* intact DNA relative to an unexcised gene *Actb*. The primers and probes used were *Vhl* forward 5′-GCTTGCGAATCCGAGGG, *Vhl* reverse 5′-TCCTCTGGACTGGCTGCC, *Vhl* Probe 5′-E6-FAM−CCCGTTCCAATAATGCCCCGG (Life Technologies) and *Actb* (mouse assay ID: Mm00607939_s1; Life Technologies). The deletion efficiency for mature B cells was 52% (95% CI 30-75%) in *Vhl*^*-/-*^*Cd19-cre* mice and 98% (95% CI, 97–99%) in *Vhl*^*-/-*^*Mb1-cre* mice(Burrows *et al*., 2020). The mice were backcrossed for at least eight generations and maintained on a C57BL/6J background. These mice, along with C57BL/6J mice (JAX, stock no. 000664) were housed in specific pathogen-free animal facilities (at 20–23 °C, with 40–60% humidity, 12-h light:12-h dark cycle). All experiments included age- and litter-matched mice that were not selected for gender. Where possible, the resource equation was used to determine sample size for experiments. Randomization was genetic and, where possible, investigators were blinded to the genetic status. For hypoxic exposure studies, a randomization algorithm was used (Excel) to allocate mice into experimental groups. Mice were immunised with 100μg 4-hydroxy-3-nitrophenylaceyl-keyhole limpet hemocyanine (NP-KLH, loading 31-33) (Biosearch Technologies) adjuvanted with Alum (Thermo Scientific) or with 0.2ml sheep red blood cells (SRBCs; packed cell volume (PCV) of 32-52%) (TCS Biosciences Ltd) via intraperitoneal injection. C57BL/6J mice were exposed to 10% O_2_ in a hypoxic chamber for 1 day, then immunised. Mice remained in the hypoxic chamber for 10 or 20 days post immunisation. Normoxic (21% O_2_) mice were treated the same way and were kept in standard conditions. For reoxygenation studies, mice were immunised and kept in the hypoxic chamber as described, on day 10 post immunisation, mice were removed from the hypoxic chamber to standard conditions for 4 or 10 days. All procedures were ethically approved by the University of Cambridge Animal Welfare and Ethical Review Body and complied with the Animals (Scientific Procedures) Act 1986 Amendment Regulations 2012, under the authority of a UK Home Office Licence. The ARRIVE (Animal Research: Reporting In Vivo Experiments) guidelines (https://arriveguidelines.org/arrive-guidelines) were used for planning, conducting and reporting experiments.

Tissue processing and immunophenotyping of murine cells by flow cytometry was performed as described in Burrows et al.(Burrows *et al*., 2020). Antibodies are listed in Supplementary table 2.

#### Murine BCR amplification and sequencing

BCR amplification and sequencing was performed as described in Burrows et al.(Burrows *et al*., 2020) Data are available at the Sequence Research Archive (SRA) database (BioProject accession nos. PRJNA574931, PRJNA574906 and PRJNA574628). Briefly, total RNA was extracted from isolated plasma cells (B220^-^CD138^+^). Reverse transcription (RT) was performed using constant region-specific primers (including unique molecular identifiers (UMIs)), followed by cDNA cleanup and PCR amplification using V gene specific primers.

Sequencing libraries were prepared using Illumina protocols and sequenced using 300bp paired-ended MiSeq (Illumina). Raw reads were filtered as Burrows et al.(Burrows *et al*., 2020) Ig gene sequence annotations were performed in IMGT V-QUEST, where somatic hypermutation repertoire and isotype usage differences were performed by custom scripts in python, and statistics were performed in *R* using Wilcoxon tests for significance (non-parametric test of differences between distributions).

#### Total Immunoglobulin (Ig) and NP-specific ELISAs

Detection of total IgM, IgG, IgA and NP-specific IgG1 was performed as Brownlie et al.(Brownlie *et al*., 2008)

#### Confocal microscopy

10µm sections were mounted on Superfrost Plus slides and air dried at RT for 1h. Samples were then fixed in -20°C acetone for 10 minutes and air dried again at RT for 1h before blocking in 0.1M Tris containing 1% BSA, 1% normal mouse serum and 1% normal rat serum. Samples were stained in a wet chamber at RT for 1h30 with the appropriate antibodies, washed 3 times in PBS and mounted in Fluoromount-G. Images were acquired using a TCS SP8 inverted confocal microscope on a 40x oil immersion objective. Raw imaging data were processed using Imaris. Antibodies are listed in Supplementary table 2.

MZ area was determined by drawing around the area of B220 positive staining (defining all B cells) and CD23 positive staining (defining the FO B cell area) within each follicle, then subtracting the CD23 positive area from the B220 area. The image analysis was performed in ImageJ.

## Data Availability

The dataset from our study can be explored interactively through a web portal: https://covid19cellatlas.org.

## Author Contributions

Conceptualization: C.H., J.R.B., P.H.M., M.R.C, J.N., P.A.L., N.B. and K.G.C.S.

Data acquisition: P.K., F.M., L.T., L.B., A.P., N.R., S.D.M., B.M.O., M.D.M., M.H and N.B.

Data Analysis: P.K., F.M., A.H., L.T., L.B., H.R., M.D.M., P.A.L. and N.B.

Project Administration: F.M., L.T., L.B., B.J.D., A.E. and C.S.

Funding Acquisition: N.K., P.J.L., C.H., J.R.B., M.P.W., P.A.L., and K.G.C.S.

Writing – Original Draft: P.J.L., C.H., P.A.L., N.B. and K.G.C.S;

Writing – Review & Editing: P.K., F.M., A.H., L.T., L.B., B.J.D., R.J.M.B., S.B., A.E., I.G.G., R.K.G., N.K., P.J.L, N.J.M., S.R., C.S., M.P.W., B.G., M.T., C.H., J.R.B, P.H.M., M.R.C., J.N., P.A.L., N.B. and K.G.C.S.

## Acknowledgements

We thank all the patients and Health Care Workers who consented to take part in this study. We are grateful for the generous support of CVC Capital Partners, the Evelyn Trust (20/75), Addenbrooke’s Charitable Trust (12/20A), the NIHR Cambridge Biomedical Research Centre and the UKRI/NIHR through the UK Coronavirus Immunology Consortium (UK-CIC) for their financial support. We thank NIHR BioResource volunteers for their participation, and gratefully acknowledge NIHR BioResource centres (Grant codes: RG85445 and RG94028), NHS Trusts and staff for their contribution. We thank the National Institute for Health Research, NHS Blood and Transplant, and Health Data Research UK as part of the Digital Innovation Hub Programme. We would like to thank: the NIHR Cambridge Clinic Research Facility outreach team for enrolment of patients; the NIHR Cambridge Biomedical Research Centre Cell Phenotyping Hub and the CRUK Cambridge Institute flow cytometry core facility for their support with flow and mass cytometry; and the Cambridge NIHR BRC Stratified Medicine Core Laboratory NGS Hub (supported by an MRC Clinical Infrastructure Award) for their support with whole blood RNA-Sequencing. The views expressed are those of the author(s) and not necessarily those of the NHS, the NIHR or the Department of Health and Social Care. K.G.C.S. is the recipient of a Wellcome Investigator Award (200871/Z/16/Z); M.P.W. is the recipient of Wellcome Senior Clinical Research Fellowship (108070/Z/15/Z); C.H. was funded by a Wellcome COVID-19 Rapid Response DCF and the Fondation Botnar; N.M. was funded by the MRC (CSF MR/P008801/1), NHSBT (WPA15-02) and Addenbrooke’s Charitable Trust (grant ref. to 900239 NJM); I.G.G. is a Wellcome Senior Fellow and was supported by funding from the Wellcome (Ref: 207498/Z/17/Z). P.J.L. is the recipient of a Wellcome Trust Principal Research Fellowship (084957/Z/08/Z) and MRC research grant (MR/V011561/1). NB and AP are supported by the Wellcome Trust, Senior Investigator Award to P.H.M, and the Rosetrees Trust. NR is supported by the NIHR Cambridge Biomedical Research Centre. Z.K.T. and M.R.C. are supported by a Medical Research Council Human Cell Atlas Research Grant (MR/S035842/1). M.R.C is supported by an NIHR Research Professorship (RP-2017-08-ST2-002). PK is the recipient of a Jacquot Research Entry Scholarship of the Royal Australasian College of Physicians Foundation.

