## Supplementary Figures for "The impact of hypoxia on B cells in COVID-19"

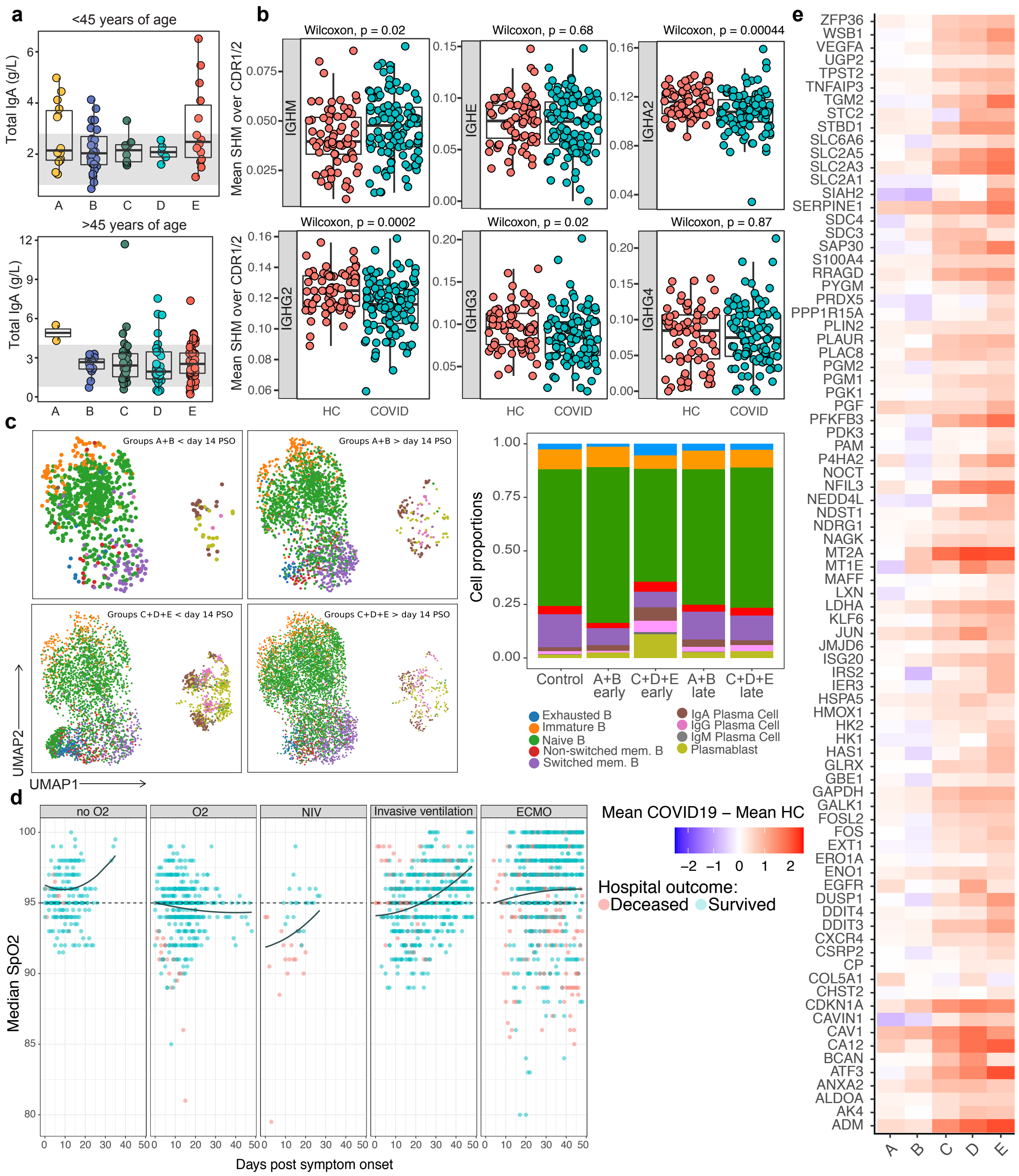


**Figure S1. B cell changes, clinical severity and hypoxia in COVID-19 patients**

**a,** Level of total IgA (g/L) detected in serum of COVID-19 cases at the time of enrolment divided into $\leq$ 45 and >45 years old age groups. Grey band correspond to 5-95$^{th}$ centile ranges based on UK Caucasian population published in the Protein Reference Unit handbook (9th Edn). **b,** Somatic hypermutation frequency calculated over the CDR1/CDR2 regions using BCR sequencing of whole blood, comparing COVID-19 cases and HCs, according to isotype, at 0-12 days post symptom onset

(Wilcoxon test FDR adjusted p-value). **c,** UMAP of B cell populations according to disease severity and days from symptom onset. Bar plot of the mean proportion of B cell populations. **d,** Median Oxygen saturations of patients with COVID according to days from symptom onset and level of oxygen supplementation. **e,** Heatmap showing gene expression for the intersection of Hypoxia GSEA leading edge genes from groups A,B,C,D and E, taken 0-12 days post screening or symptom onset. **a,b,d,** Circles represent individual donors.

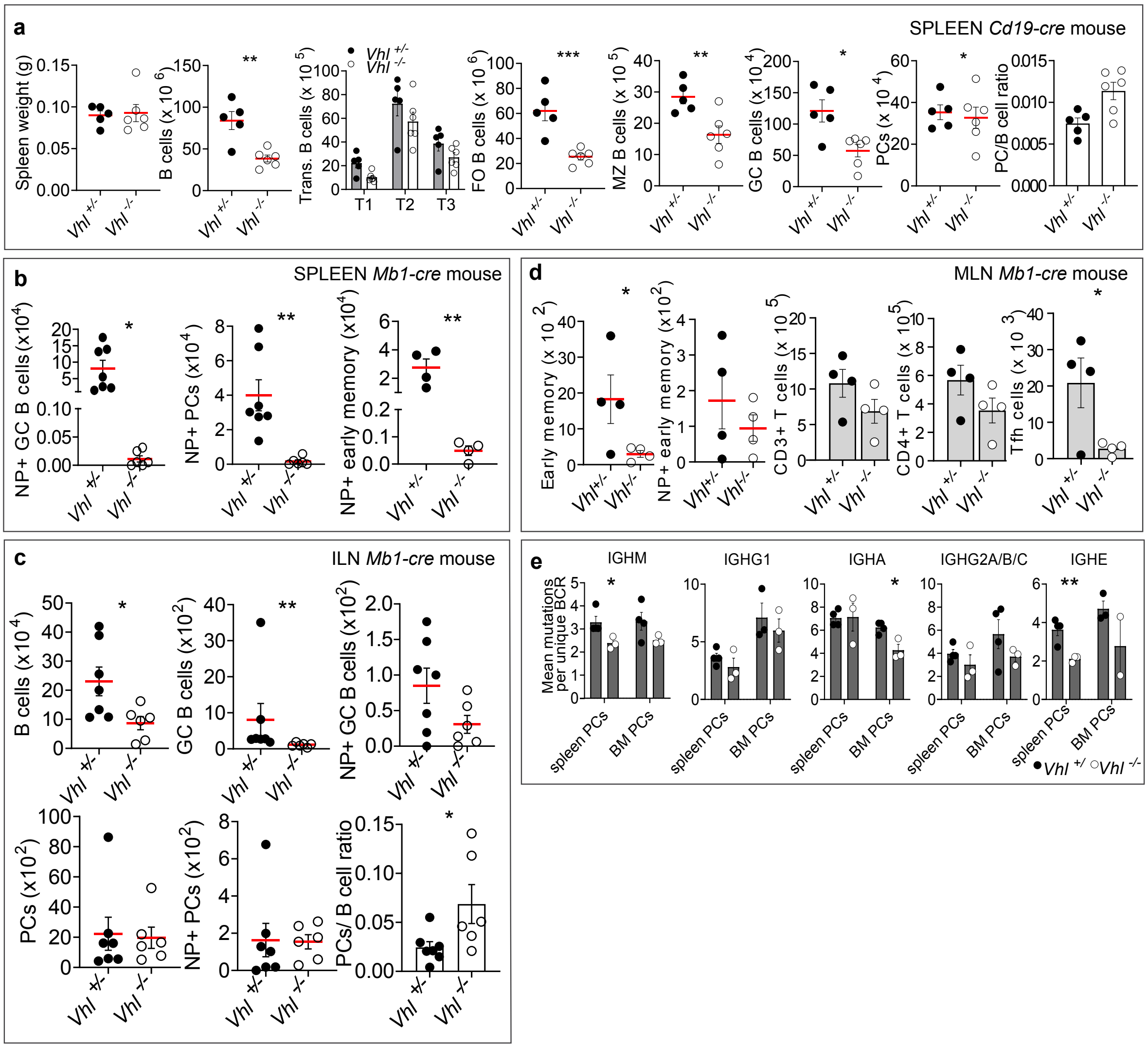


**Figure S2. Constitutive HIF activation in mice leads to reduced antigen-specific B cells, Tfh cells and SHM.**

**a,** B cell flow cytometric data from *Vhl*$^{+/-}$*Cd19-cre* and *Vhl*$^{-/-}$*Cd19-cre* mice 10 days post sheep red blood cell (SRBC) immunization; Spleen B cells, Transitional, FO, MZ, GC B cells and PCs. **P* < 0.05,***P* < 0.01,****P* < 0.001 unpaired, two-sided Student's t-test. B cell flow cytometric data from *Vhl*$^{+/-}$*Mb1-cre* and *Vhl*$^{-/-}$*Mb1-cre* mice 21 days post NP-KLH immunization in **b,** Spleen; NP-specific GC, PC and early memory B cells. **P* < 0.05,***P* < 0.01, ****P* < 0.001 , *****P* < 0.0001 unpaired, two-sided Student's t-test. **c,** ILN (inguinal lymph node); total, GC, NP+ GC, PC, NP+ PC and PC:B cell ratio displayed. PCs were not increased in absolute number, but the PC:B cell ratio was consistently increased. **P* < 0.05 unpaired, two-sided Student's t-test; GC B cells ***P* < 0.01 two-sided Mann-Whitney test. **d,** MLN (mesenteric lymph node) total and NP+ early memory B cells, T cells ($CD3^+$ and $CD4^+$) and T follicular helper (Tfh) cells ($CD3^+CD4^+PD\text{-}1^{high}CXCR5^{high}FoxP3^-$), displayed. early memory; *$P < 0.05$ unpaired, two-sided Student's t-test, Tfh; *$P < 0.05$ unpaired, two-sided Student's t-test. **e,** Mean base-pair mutations per unique BCR per isotype (relative to reference germline IGHV gene) in naïve *Vhl*$^{+/-}$*Mb1-cre* and *Vhl*$^{-/-}$*Mb1-cre* mice. *$P < 0.05$,**$P < 0.01$ unpaired, two-sided Student's t-test. **(a-e)** Gated as in Fig. 1, mean ± s.e.m, individual mice shown. Early memory, T cell and data in **e,** from one experiment, all other data are pooled from two independent experiments.

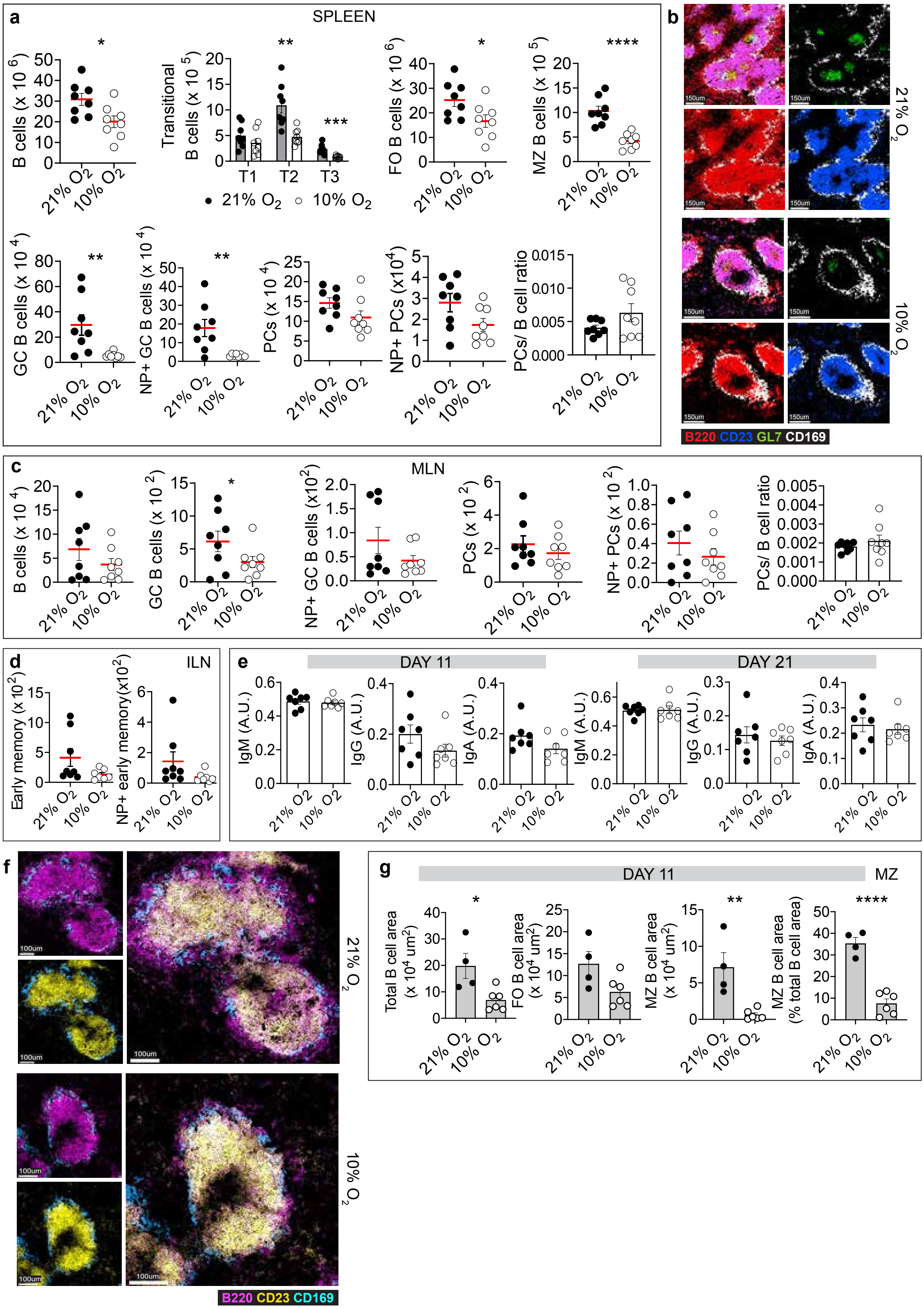


**Figure S3.** ***In vivo*** **hypoxia leads to marked and persistent B cell defects in mice.**

**a,** WT mice were exposed to 21% $O_2$ (normoxia) or 10% $O_2$ (hypoxia), were immunized with NP-KLH at d 1, then at d 11 splenic B cell subsets were enumerated by flow cytometry. Total, transitional, FO, MZ, GC, NP+ GC B cells, PCs, NP+ PCs and PC: B cell ratio is displayed. *$P < 0.05$,**$P < 0.01$,***$P < 0.001$,****$P < 0.0001$ unpaired, two-sided Student's t-test, spleen. **b,** Representative spleen confocal images from mice immunised as in (Fig. 4a) and harvested on d 11. GC B cells (green, $B220^+GL7^+$), B cells (red, $B220^+$), FO B cells (blue, $B220^+CD23^+$) and MZ metallophilic macrophages (white, $CD169^+$). WT mice were exposed to 21% $O_2$ (normoxia) or 10% $O_2$ (hypoxia), were immunized with NP-KLH at d 1, then at d 21 B cell subsets were enumerated by flow cytometry in **c,** MLN; total, GC, NP+ GC, PC, NP+ PC and PC:B cell ratio displayed. *$P < 0.05$ unpaired, two-sided Student's t-test, **d,** ILN; total and NP+ early memory B cells displayed. Unpaired, two-sided Student's t-test. **e,** Serum IgM, IgG and IgA in normoxic and hypoxic mice immunised with NP-KLH on d 1 then harvested on d 11 or 21, by ELISA (A.U., arbitrary units). Unpaired, two-sided Student's t-test. **f,** Representative spleen confocal images from mice immunised as in (Fig. 4a) and harvested on d 11. MZ B cells (pink, $B220^+CD23^-$), FO B cells (yellow, $B220^+CD23^+$) and MZ metallophillic macrophages (blue, $CD169^+$). **g,** B cell and MZ area in spleens from immunised mice described in Fig. 4a and harvested on d 11. Symbols represent individual follicles from one spleen per condition. *$P < 0.05$,**$P < 0.01$ ,****$P < 0.01$ unpaired, two-sided Student's t-test. **(a-d,f)** $n$ = 8 21% $O_2$ and 8 10% $O_2$, **(e)** $n$ = 7 21% $O_2$, 7 10% $O_2$, individual mice. **(a-d,f)** Data pooled from two independent experiments, results confirmed in a third. **(e)** Data, represents three independent experiments. **(a-g)** Mean ± s.e.m.

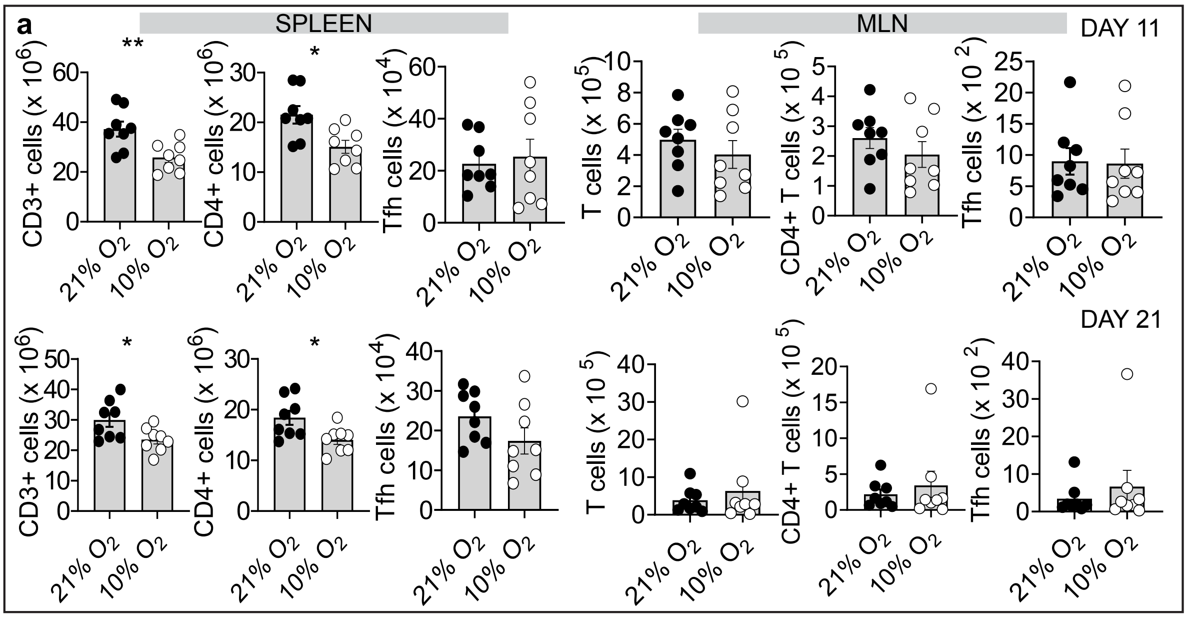


**Figure S4.** ***In vivo*** **hypoxia has minor effects on T cells after 11 or 21 day exposures in mice**

T cells and Tfh cells from normoxic and hypoxic mice immunised with NP-KLH on d 1 then harvested on d 11 or 21 (gated as in S. 2d). $*P < 0.05$,$**P < 0.01$ unpaired, two-sided Student's t-test. $n$ = 8 21% $O_2$ and 8 10% $O_2$. Data pooled from two independent experiments. Mean ± s.e.m.
